# Rethinking under-vaccination: social identity and its association with vaccination attitudes and hesitancy-related behaviour

**DOI:** 10.64898/2026.05.25.26354049

**Authors:** Leah Borovoi, Rotem Kahalon, Michael Edelstein

**Affiliations:** Azrieli Faculty of Medicine, Bar-Ilan University, Safed, Israel

**Keywords:** under-vaccination, identity centrality, social identity, online affiliations, risk stratification, vaccine hesitancy

## Abstract

Research on under-vaccination often segments populations using demographic or administrative variables that are operationally useful but fail to capture identity dimensions relevant to vaccination decisions. Drawing on social identity theory, we propose an identity-landscape approach distinguishing identity membership, identity centrality, and multidimensional identity structure. Using a cross-sectional survey of 1,000 UK parents, we measured 65 identity indicators, identity-importance ratings, and their association with attitudinal and behavioural hesitancy toward childhood vaccination using validated scales. Beyond established socio-demographic predictors, alternative-medicine and natural-lifestyle identities, as well as affiliation with social media networks, were linked to greater hesitancy. Greater centrality of religion and political affiliation within personal identity was also associated with higher hesitancy. Principal component analysis suggested that individuals actively engaged across multiple societal issues were more hesitant, whereas stereotypically male-gendered engagement was associated with lower hesitancy. An identity-focused population segmentation may identify previously unrecognized undervaccinated groups and inform innovative tailored immunization campaigns.

## BACKGROUND

Controlling vaccine-preventable diseases depend on sustained uptake across the population. While routine childhood programmes are universal in their nature, under-vaccination, understood as delayed, incomplete, or absent vaccination, is well recognised across immunisation programmes, with some subgroups having lower vaccination uptake than the wider population from which they originate (18). Health policy has at times attributed under-vaccination to socioeconomic deprivation or health-system failure alone, but although this factor is a key contributor, undervaccination is the result of a complex interplay between systems, populations and individual factors that interact differently in different population groups (18, 19, 25). In practice, public health services often identify under-vaccinated populations using a small set of routinely available markers such as age, deprivation, geography, ethnicity, religion, or administrative status, interprets the resulting associations as if they captured relevant identity processes (3, 24, 25, 51) and then design vaccine campaigns around these labels (12, 13, 24, 25). This pragmatic approach is often necessary for delivery. However, these variables are used not because they fully explain variation in vaccination, but because they are available in standard public-health datasets.

Such an approach, which relies solely on routine dataset variables, has at least three limitations. First, category membership does not tell us how salient a given identity is to a given individual (i.e., a person can be categorised as belonging to a specific ethnic group, yet the importance of belonging to this group to the person’s perceived identity can vary). Second, standard models usually consider one identity dimension at a time, rather than conceptualising identity as a constellation of identities that coexist and interact within the same person. Third, routine datasets often miss other potential drivers of vaccine attitudes that operate through sociocultural affiliations and information environments, including lifestyle movements, political leanings, professional and academic networks, and online affiliations, which, if relevant to one’s identity, may shape norms, trust, and exposure to vaccine-related narratives. These dimensions require direct measurement of how people describe and value their own identities.

We use social identity theory and multidimensional models of collective identity to examine under-vaccination through a broader account of identity that includes identity category membership (whether individuals self-identify as members of a given social category)category) and identity centrality (the extent to which this category membership is personally meaningful and central to their self-concept).), and structure (the interplay of multiple co-existing identities within the same person) (2, 44, 46, 48). Rather than replacing conventional epidemiological stratification, this approach adds a psychological layer by distinguishing identity category membership, identity centrality, and multidimensional identity structure.

Identity category membership refers to whether a person endorses belonging to a given social category or group, for example, a religion, profession, lifestyle movement, or online affiliation. Social identity theory treats social categories and roles as important bases of self-definition, so membership is not trivial (44, 46). This category membership can represent the outcome of several concurrent mechanisms (e.g., social norms, service design, discrimination, access barriers, or historical distrust). A regression coefficient for “religion” or “migrant background” does not tell us which of this mechanism is impacting the outcome of interest. The association between the identity captured by the category membership and the outcome is a combination of mechanisms that traditional epidemiological association studies cannot disentangle.

Identity centrality refers to the degree to which a particular identity is central to an individual’s self-perception. Consistent with the social identity approach, the psychological importance of group membership often predicts attitudes, well-being, and behaviour beyond category membership alone (10, 14, 31, 45). Two people may both belong to the same group, yet the group may be self-defining for only one of them; in that case, group norms, trusted messengers, and motivated reasoning may plausibly be more influential for the person for whom the identity is more central. For example, stronger gender identification has been linked to greater collective action around gender inequality (4) and in a recent meta-analysis positive association was found between social identity and health-related behavior (14).

Finally, within social identity theory, the self is understood as comprising multiple social identities that vary in their centrality and salience across contexts (31, 48). Individuals do not carry one identity at a time; they occupy overlapping roles and memberships, including parenthood, profession, religion, migration history, sexuality, lifestyle, political orientation, neighbourhood, and digital affiliations (6). The organisation of these identities, referred to here as identity structure, has been conceptualised in terms of the complexity and configuration of individuals’ multiple group memberships (41). For public health, this suggests that risk may be attached not only to single labels but to recurring configurations or bundles of identities. Taken together, the distribution of identity category membership, identity centrality, and recurring combinations of identities within a population form the identity landscape.

Evidence from COVID-19 research supports this broader view that factors beyond socio-demographic variables impacted on vaccination attitudes and behaviour: during the pandemic, factors such as adherence to collective norms and identification with national identity were associated with support for public-health measures (49, 50). Identity is not a deterministic explanation of vaccination behaviour. It is a measurable route through which people may interpret risk, decide whom to trust, and respond to vaccination programmes. Online affiliation is increasingly part of this complex identity structure. In addition to information, online platforms provide community cues, reputation systems, algorithmic curation, and repeated normative feedback. Such online information patterns can be framed as population signals relevant to public health (22, 23). Online affiliations may also make identities more salient. Recent research applying social identity theory to digital contexts argues that anonymity, group structure, and feedback mechanisms can reinforce identity-consistent norms and behaviours (6).

Drawing on social identity theory, we examined which social identities, online and offline, are most strongly associated with vaccine attitudes and behaviour, based on the premise that identities influence attitudes and behaviour through norms and shared meanings. The objective was to assess whether identity category membership and importance were associated with parental attitudinal or behavioural hesitancy toward routine childhood vaccination, using a cross-sectional survey.

## METHODS

### Survey design

We developed the survey iteratively, combining standard sociodemographic items with identity domains that are not usually available in routine public-health data, using questions and scales from a range of published and standardized tools, described below. Although the study purported to examine routine childhood vaccination in general, the survey explicitly cited the measles-mumps-rubella (MMR) and diphtheria-tetanus-pertussis-polio-Hib-hepatitis B (6-in-1) vaccines as examples.

For identity category membership, we used or adapted standard classifications and categories from the 2021 England National census for ethnicity, religion, educational attainment, sexual orientation, and occupation (34). Place of residence used a simplified rural/small town/urban classification informed by UK rural-urban classification frameworks (15). National, global, and political identity-related questions were developed and adapted from the European Social Survey (21). Questions relating to social and community identity were drawn from the in-group identification scale (31). Other domains, including lifestyle identities, online affiliations, academic affiliations, community roles, cultural-interest groups, were study-developed or theory-informed (2, 6, 44, 46, 48) to capture self-chosen and socially embedded identities not routinely represented in health datasets or in the literature.

Participants also rated the extent to which the relevant identity was central to their self-concept using one item: “My [XX] is an important part of who I am.” This item was adapted from the centrality component of Leach et al.’s (31) multidimensional model of in-group identification. We used 1-7 Likert scales dichotomised as high (5–7) versus low (<5).

The main attitudinal outcome was the 4-item short form of the Parent Attitudes about Childhood Vaccines survey (PACV-SF) (35, 36). Each of the four items (concern about vaccine effectiveness, overall hesitancy, trust in vaccine information, and prior delay of a routine vaccine for reasons other than illness or allergy) was coded 0 or 1 using predefined hesitant-response definitions, yielding a raw score of 0-4. For comparability with the conventional PACV metric, scores were linearly transformed to a 0-100 scale; attitudinal hesitancy was defined as a transformed score >50 (equivalent to a raw score of 3-4). Behavioural hesitancy was coded as present if respondents reported ever delaying a routine childhood vaccine for reasons other than illness or allergy, vaccinating their child(ren) with some but not all recommended vaccines, or usually not vaccinating their child(ren). Two instructed-response attention checks were embedded to assess response quality. To reduce demand characteristics and limit awareness of the hypothesis, the survey included a parallel block of items on antibiotic use, presented alongside vaccine-related questions.

The questionnaire was piloted with 10 participants and modified to its final version based on their feedback. The analysed domains are summarised in Appendix 1.

Parents of children under the age of 10 residing in the United Kingdom were recruited between 23 and 27 November 2025 through Prolific, an online participant-recruitment platform widely used in behavioural research (16, 39).

The questionnaire began with consent and parental eligibility screening. Respondents reported the number of children and the age of their youngest child; those with no children or with all children older than 10 years were excluded.

The target sample size of approximately 1,000 participants was selected pragmatically based on several analytic considerations rather than a single hypothesis-testing calculation. The study aimed to examine a large number of identity indicators and their covariance structure, including exploratory PCA of approximately 65 identity-related variables, for which common recommendations suggest participant-to-variable ratios of approximately 10:1 to 15:1. The sample size was also intended to support stable subgroup comparisons and adjusted regression analyses across multiple identity domains while reducing sparse-cell instability and allowing for quality-control exclusions.

The study received approval from the Bar-Ilan University Faculty of Medicine Ethics Committee.

### Analysis

We first described the distribution of each variable and estimated outcome prevalence using means and proportions. To reduce sparse-cell instability and ensure adequate comparison-group sizes, sub-categories where merged where necessary, for example Ethnicity was initially measured using detailed UK census-style categories and was collapsed for analysis into White, Black, Asian, and Mixed/Other groups. Likewise, religious groups other than Christian and Muslim were merged into one “other” group. For each identity indicator and centrality measure we calculated unadjusted risk ratios using modified Poisson regression with robust standard errors. Adjusted estimates were calculated for exposures significantly associated with one of our two outcomes at the alpha=0.05 level.

To examine the multidimensional structure of identity, we used principal component analysis (PCA), a dimension-reduction method that summarises correlated patterns among variables into components (1). PCA was conducted in Python using scikit-learn (sklearn.decomposition.PCA), with prior standardisation of all variables (z-scores). Components were extracted by singular value decomposition without rotation from the identity-only matrix of 65 derived binary indicators. The feature set included binary indicators for mutually exclusive identity categories and binary checklist indicators, but excluded age, socioeconomic variables, family-structure variables, identity-importance variables, and the outcomes themselves. For each component, outcome associations were estimated per 1-standard-deviation increase in the standardised component score, using the same modelling framework and adjustment set.

Primary outcomes and PCA scores were complete for all respondents. Centrality was analysed only among respondents for whom the relevant identity-importance question was applicable and asked; structurally absent responses (arising from survey routing logic) were excluded rather than recoded as low importance. Model-specific denominators are reported in Supplementary Tables S1-S4.

Given the large number of identity indicators examined, p-values from the association models were adjusted for multiple testing using the Benjamini-Hochberg false discovery rate (FDR) procedure. Correction was applied separately within each family of analyses (identity membership and identity centrality models).

## RESULTS

### Characteristics of the sample

Of the 1,000 recruited participants, 601 (60.1%) identified as women, 414 (41.4%) reported living in an urban area, 465 (46.5%) reported having two children, 665 (66.5%) reported higher education, and 235 (23.5%) reported household income above £80,000 (Table 1). Overall, 37.3% reported adhering to a particular lifestyle, 52.3% reported belonging to an online community, 57.1% reported engagement with a particular cause, and 25.1% reported a community role (Table 1). Attitudinal hesitancy (PACV

**Table 1.**
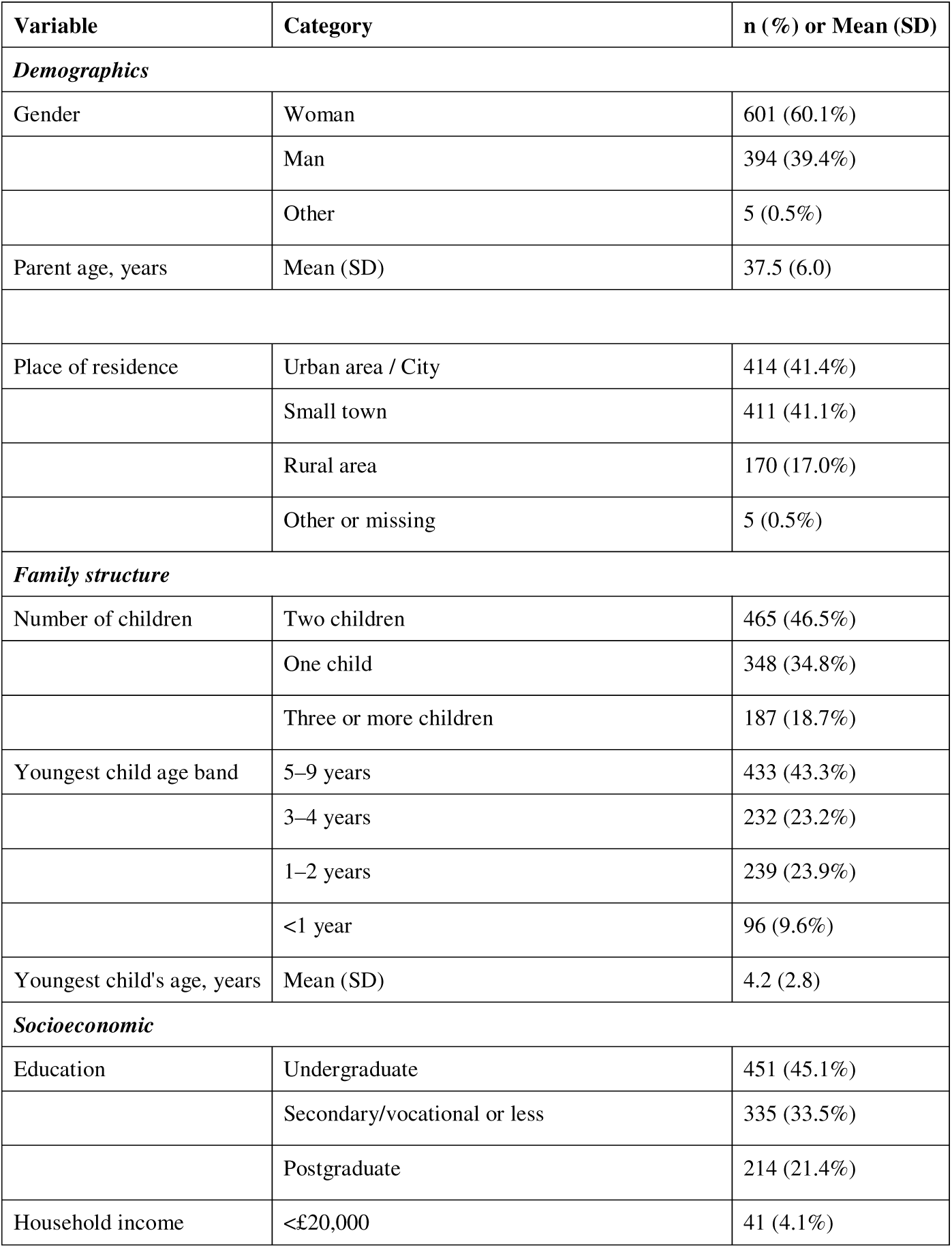
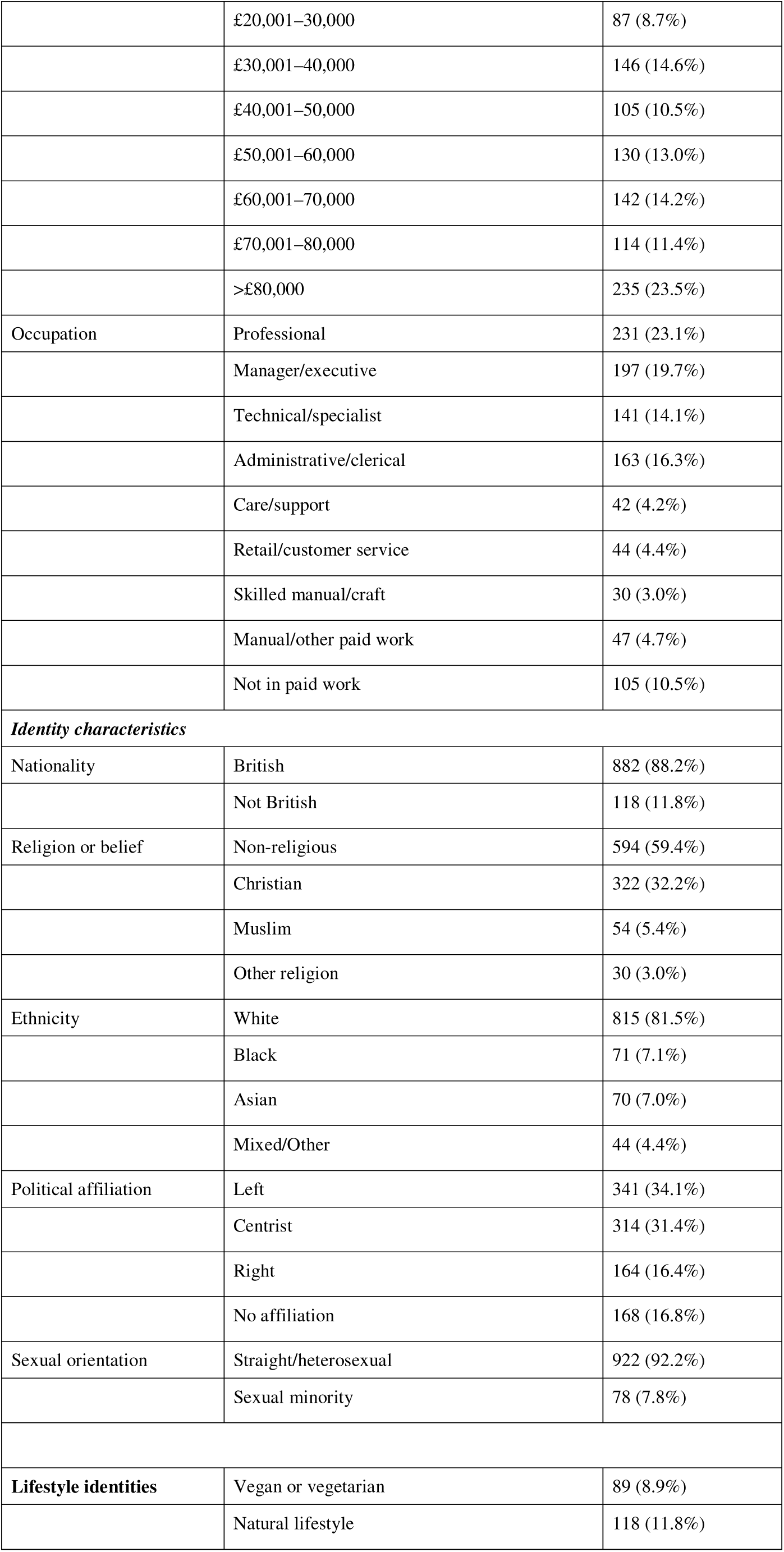
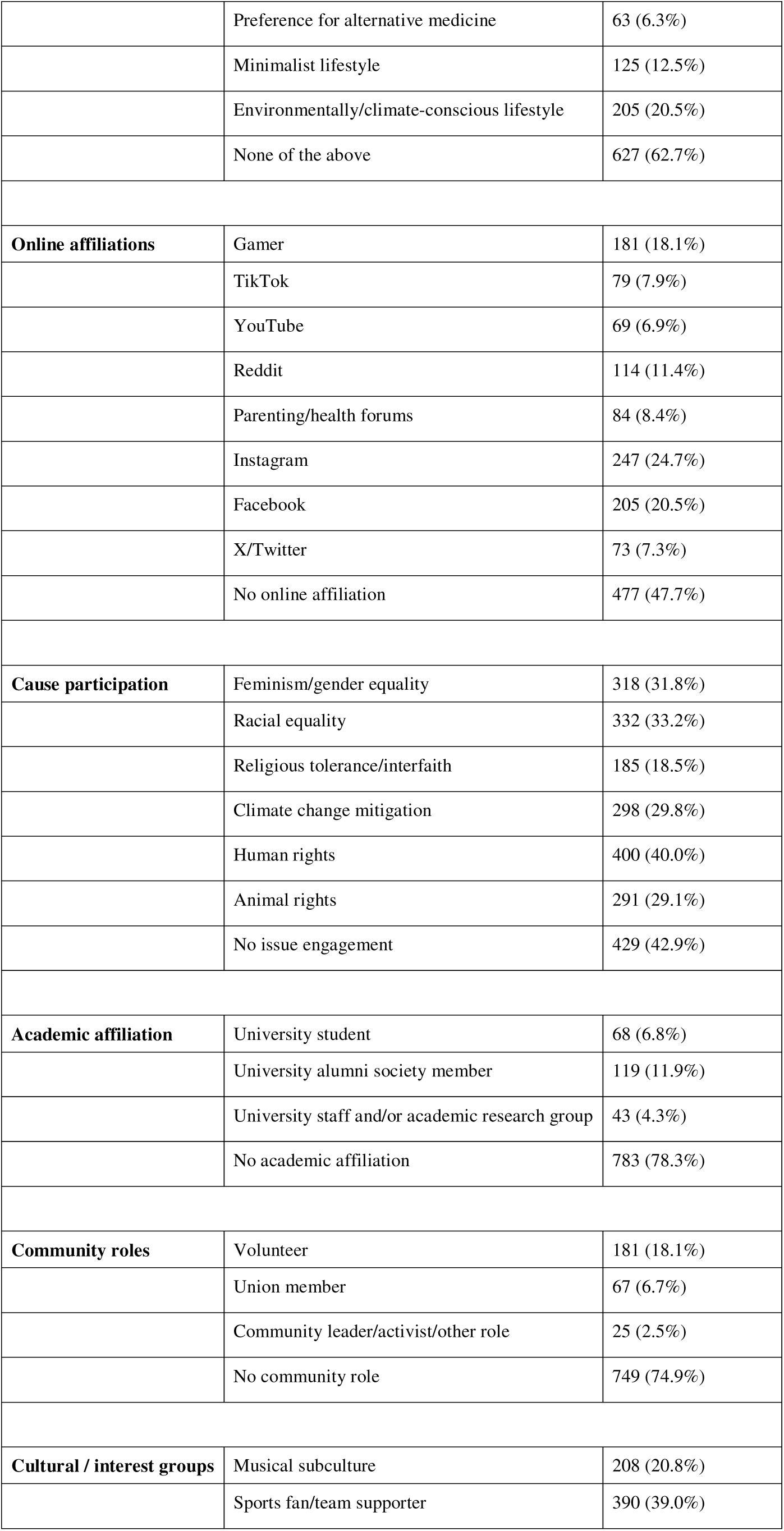
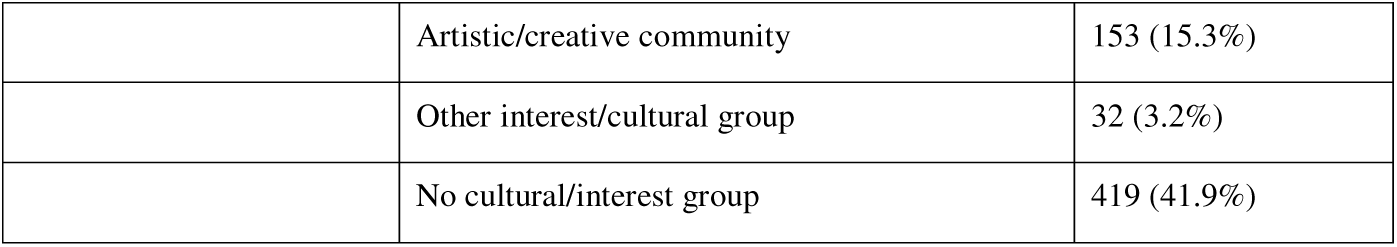
Characteristics of the analytic sample (n=1000)

| Variable | Category | n (%) or Mean (SD) |
| --- | --- | --- |
| **Demographics** |  |  |
| Gender | Woman | 601 (60.1%) |
|  | Man | 394 (39.4%) |
|  | Other | 5 (0.5%) |
| Parent age, years | Mean (SD) | 37.5 (6.0) |
| Place of residence | Urban area / City | 414 (41.4%) |
|  | Small town | 411 (41.1%) |
|  | Rural area | 170 (17.0%) |
|  | Other or missing | 5 (0.5%) |
| **Family structure** |  |  |
| Number of children | Two children | 465 (46.5%) |
|  | One child | 348 (34.8%) |
|  | Three or more children | 187 (18.7%) |
| Youngest child age band | 5–9 years | 433 (43.3%) |
|  | 3–4 years | 232 (23.2%) |
|  | 1–2 years | 239 (23.9%) |
|  | <1 year | 96 (9.6%) |
| Youngest child's age, years | Mean (SD) | 4.2 (2.8) |
| **Socioeconomic** |  |  |
| Education | Undergraduate | 451 (45.1%) |
|  | Secondary/vocational or less | 335 (33.5%) |
|  | Postgraduate | 214 (21.4%) |
| Household income | <£20,000 | 41 (4.1%) |
|  | £20,001–30,000 | 87 (8.7%) |
|  | £30,001–40,000 | 146 (14.6%) |
|  | £40,001–50,000 | 105 (10.5%) |
|  | £50,001–60,000 | 130 (13.0%) |
|  | £60,001–70,000 | 142 (14.2%) |
|  | £70,001–80,000 | 114 (11.4%) |
|  | >£80,000 | 235 (23.5%) |
| Occupation | Professional | 231 (23.1%) |
|  | Manager/executive | 197 (19.7%) |
|  | Technical/specialist | 141 (14.1%) |
|  | Administrative/clerical | 163 (16.3%) |
|  | Care/support | 42 (4.2%) |
|  | Retail/customer service | 44 (4.4%) |
|  | Skilled manual/craft | 30 (3.0%) |
|  | Manual/other paid work | 47 (4.7%) |
|  | Not in paid work | 105 (10.5%) |
| **Identity characteristics** |  |  |
| Nationality | British | 882 (88.2%) |
|  | Not British | 118 (11.8%) |
| Religion or belief | Non-religious | 594 (59.4%) |
|  | Christian | 322 (32.2%) |
|  | Muslim | 54 (5.4%) |
|  | Other religion | 30 (3.0%) |
| Ethnicity | White | 815 (81.5%) |
|  | Black | 71 (7.1%) |
|  | Asian | 70 (7.0%) |
|  | Mixed/Other | 44 (4.4%) |
| Political affiliation | Left | 341 (34.1%) |
|  | Centrist | 314 (31.4%) |
|  | Right | 164 (16.4%) |
|  | No affiliation | 168 (16.8%) |
| Sexual orientation | Straight/heterosexual | 922 (92.2%) |
|  | Sexual minority | 78 (7.8%) |
| **Lifestyle identities** | Vegan or vegetarian | 89 (8.9%) |
|  | Natural lifestyle | 118 (11.8%) |
|  | Preference for alternative medicine | 63 (6.3%) |
|  | Minimalist lifestyle | 125 (12.5%) |
|  | Environmentally/climate-conscious lifestyle | 205 (20.5%) |
|  | None of the above | 627 (62.7%) |
| **Online affiliations** | Gamer | 181 (18.1%) |
|  | TikTok | 79 (7.9%) |
|  | YouTube | 69 (6.9%) |
|  | Reddit | 114 (11.4%) |
|  | Parenting/health forums | 84 (8.4%) |
|  | Instagram | 247 (24.7%) |
|  | Facebook | 205 (20.5%) |
|  | X/Twitter | 73 (7.3%) |
|  | No online affiliation | 477 (47.7%) |
| **Cause participation** | Feminism/gender equality | 318 (31.8%) |
|  | Racial equality | 332 (33.2%) |
|  | Religious tolerance/interfaith | 185 (18.5%) |
|  | Climate change mitigation | 298 (29.8%) |
|  | Human rights | 400 (40.0%) |
|  | Animal rights | 291 (29.1%) |
|  | No issue engagement | 429 (42.9%) |
| **Academic affiliation** | University student | 68 (6.8%) |
|  | University alumni society member | 119 (11.9%) |
|  | University staff and/or academic research group | 43 (4.3%) |
|  | No academic affiliation | 783 (78.3%) |
| **Community roles** | Volunteer | 181 (18.1%) |
|  | Union member | 67 (6.7%) |
|  | Community leader/activist/other role | 25 (2.5%) |
|  | No community role | 749 (74.9%) |
| **Cultural / interest groups** | Musical subculture | 208 (20.8%) |
|  | Sports fan/team supporter | 390 (39.0%) |
|  | Artistic/creative community | 153 (15.3%) |
|  | Other interest/cultural group | 32 (3.2%) |
|  | No cultural/interest group | 419 (41.9%) |

### Associations with self-identified memberships and affiliations

Table 2 presents associations that remained statistically significant after adjustment. The full membership and centrality results (crude and adjusted) are reported in Supplementary Tables S1-S4. Several identity indicators were associated with one or both hesitancy outcomes. Compared with the highest-income group (>£80,000), the lowest-income group (<£20,000) had markedly higher estimated risk of attitudinal hesitancy (aRR 3.92; 95% CI 2.49-6.16) and behavioural hesitancy (aRR 3.38; 95% CI 1.43-8.03). Identifying as Black or Asian was associated with higher attitusinal hesitancy compared to identifying as White (aRR 2.30 [1.59, 3.31] and 1.64 [1.12, 2.42] respectively). Identifying with being Christian or Muslim was also associated with higher attitudinal hesitancy (aRR 1.58 [1.21, 2.05] and 1.89 [1.28, 2.79] respectively), whereas identifying as being political affiliated with the left was associated with less hesitancy (Table 2). Self-identification with specific lifestyle movements showed similarly large associations: alternative-medicine identity was associated with attitudinal hesitancy (aRR 3.17; 95% CI 2.40-4.18) and behavioural hesitancy (aRR 3.98; 95% CI 2.35-6.75), and natural-lifestyle identity was associated with attitudinal hesitancy (aRR 2.33; 95% CI 1.80-3.03) and behavioural hesitancy (aRR 2.73; 95% CI 1.68-4.45).

**Table 2.** Selected adjusted associations in the UK parent case example.

| Domain | Variable or contrast | Attitudinal hesitancy, PACV >50, aRR [95% CI] | Behavioural hesitancy, delay/refusal, aRR [95% CI] |
| --- | --- | --- | --- |
| Place of residence | Small town (n=411) vs City (n=414) | 0.74 [0.57-0.98] | 0.56 [0.34 - 0.92] |
| Socioeconomic position | Income <£20,000 (n=41) vs. >£80,000 (n=235) | 3.92 [2.49, 6.16] | 3.38 [1.43, 8.03] |
|  | Income <£20,001-30,000 (n=87) vs. >£80,000 (n=235) | 2.27 [1.43, 3.61] |  |
|  | Income <£30,001-40,000 (n=146) vs. >£80,000 (n=235) | 1.83 [1.2, 2.78] |  |
| Religion or belief | Christian (n=322) vs. non-religious (n=594) | 1.58 [1.21, 2.05] | 1.96 [1.25, 3.07] |
|  | Muslim (n=54) vs. non-religious (n=594) | 1.89 [1.28, 2.79] | — |
|  | Other religion (n=30) vs. non-religious (n=594) | 2.52 [1.60, 3.97] | — |
| Ethnicity | Black vs. White | 2.30 [1.59, 3.31] | - |
|  | Asian vs. White | 1.64 [1.12, 2.42] | - |
|  | Mixed/Other vs. White | 1.82 [1.14, 2.89] | — |
| Political affiliation | Centrist (n=314) vs. Left (n=341) | 1.41 [1.01, 1.97] | — |
|  | Right (n=164) vs. Left (n=341) | 1.89 [1.33, 2.69] | — |
|  | No political affiliation (n=168) vs. Left (n=341) | 1.88 [1.30, 2.72] | — |
| Lifestyle and community identity (vs those not identify with the particular lifestyle) | Alternative-medicine identity (n=63) | 3.17 [2.40, 4.18] | 3.98 [2.35, 6.75] |
|  | Natural-lifestyle identity (n=118) | 2.33 [1.80, 3.03] | 2.73 [1.68, 4.45] |
|  | Minimalist lifestyle (n=125) | 1.52 [1.15, 2.01] | - |
|  | Community leader/activist | 1.64 [1.06, 2.54] | 2.28 [1.06, 4.88] |
|  | No lifestyle identity (n=627) | 0.57 [0.45, 0.73] | 0.53 [0.34, 0.82] |
| Online affiliation (vs those not identify with the particular online community) | TikTok affiliation (n=79) | 1.38 [0.99, 1.93] | 2.24 [1.33, 3.77] |
|  | YouTube affiliation (n=69) | 1.73 [1.23, 2.45] | 2.02 [1.14, 3.58] |
|  | Instagram affiliation | 1.51 [1.18, 1.94] | 1.57 [1.03, 2.39] |
|  | X/Twitter affiliation | 1.95 [1.38, 2.75] | 2.19 [1.17, 4.10] |
|  | No online affiliation | 0.69 [0.54, 0.88] | — |
| Identity centrality | High religion centrality vs. low | 2.20 [1.43, 3.37] | - |
|  | High political-affiliation centrality vs. low | — | 1.70 [1.04, 2.77] |
|  | High “not engaging” centrality vs. low | 2.61 [1.81, 3.78] | 3.18 [1.46, 6.92] |
|  | High sexual-orientation centrality vs. low | 1.56 [1.15, 2.10] | 2.03 [1.15, 3.60] |
| PCA identity structure | PC1: engagement–disengagement axis, per 1 SD | 1.27 [1.14, 1.42] | 1.35 [1.13, 1.61] |
|  | PC2: gendered/digital participation axis, per 1 SD | 0.80 [0.70, 0.91] | - |

**Abbreviations:** aRR, adjusted risk ratio; CI, confidence interval; --, not statistically significant at conventional thresholds in the case example models.

Self-identified affiliation with YouTube, Instagram or X/Twitter were associated with increased risk of attitudinal hesitancy compared to no use (aRR 1.73, 1.51 and 1.95 respectively, table 2), whereas reporting no self-identified online affiliation was associated with lower risk (aRR 0.69; 95% CI 0.54-0.88). Behavioural hesitancy was associated with Affiliation with TikTok (aRR 2.24; 95% CI 1.33-3.77), YouTube (aRR 2.02; 95% CI 1.14-3.58), Instagram (aRR 1.57; 95% CI 1.03-2.39), and X/Twitter (aRR 2.19; 95% CI 1.17-4.10).

### Centrality beyond membership

Sexual orientation clearly illustrated the importance of centrality vs membership: in a model including both membership and centrality, sexual-minority membership was not positively associated with attitudinal hesitancy (aRR 0.66; 95% CI 0.44-1.00), whereas high sexual-orientation centrality was associated with higher attitudinal hesitancy (aRR 1.56; 95% CI 1.15-2.10) and behavioural hesitancy (aRR 2.03; 95% CI 1.15-3.60). High religion centrality (i.e considering religion as central to one’s identity regardless of religious affiliation) was associated with attitudinal hesitancy (aRR 2.20; 95% CI 1.43-3.37) whereas the centrality of political affiliation (regardless of political leanings) was associated with behavioural hesitancy (aRR 1.70 [1.04, 2.77]). While not engaging in a particular lifestyle or online community was associated with lower hesitancy, those who indicated that not being engaged was a central element to their identity was associated with higher attitudinal hesitancy (aRR 2.61; 95% CI 1.81-3.78) and behavioural hesitancy (aRR 3.18; 95% CI 1.46-6.92, Table 2, Supplementary Tables S2 and S4).

### Identity structure (PCA)

PCA identified three interpretable components (PC1, PC2, and PC3) explaining 9.62%, 6.06%, and 5.98% of the variance in the 65 identity indicators, respectively. PC1 summarised differences in engaged versus disengaged identity participation across the issue-engagement, lifestyle, academic, community, and digital domains. Positive loadings were strongest for human rights, climate change mitigation, racial equality, environmentally/climate-conscious lifestyle, and religious tolerance/interfaith; negative loadings were strongest for repeated none categories, especially no issue engagement, no lifestyle identity, no academic affiliation, no community role, and no online affiliation. Higher PC1 was associated with attitudinal hesitancy (aRR 1.27; 95% CI 1.14-1.42) and behavioural hesitancy (aRR 1.35; 95% CI 1.13-1.61).

PC2 captured a gendered pattern of participation and online engagement. Positive loadings were strongest for male gender, sports fandom, X/Twitter affiliation, gaming affiliation, YouTube affiliation, straight/heterosexual identity, and crypto affiliation; negative loadings were strongest for woman, feminism/gender equality, no online affiliation, no cultural affiliation, racial equality, animal rights, sexual-minority identity, and vegan/vegetarian identity. Higher PC2 was inversely associated with attitudinal hesitancy (aRR 0.80; 95% CI 0.70-0.91) and was not clearly associated with behavioural hesitancy (aRR 0.88; 95% CI 0.68-1.13).

PC3 captured a nationality-ethno-religious covariance pattern: dominant positive loadings reflected non-British nationality, Black ethnicity, and Muslim identity, whereas dominant negative loadings reflected British nationality, White ethnicity, and non-religious identity. PC3 was not clearly associated with either outcome (attitudinal aRR 0.97; 95% CI 0.84-1.11; behavioural aRR 1.13; 95% CI 0.89-1.43) (Supplementary Table S5). A null association for PC3 does not imply that all variables contributing to that component are individually unrelated to hesitancy. PCA captures shared covariance structure, and a component can therefore be unrelated to the outcome even when some constituent variables show one-at-a-time associations.

## DISCUSSION

This study expands the well-established concept of socio-demographic predictors of vaccination behaviour by deconstructing group membership into membership and centrality, exploring previously understudied predictors, and taking into account the complexity of multi-layered identity structures. Our findings are consistent with established evidence that under-vaccination is heterogeneous and patterned by socioeconomic position, ethnicity, and religion (7, 12, 17, 19, 24, 33). They also fit with work showing that vaccine hesitancy reflects psychological antecedents, such as confidence and collective responsibility, as well as broader contextual and social influences (5, 30). In this case, the lower income and several ethnicity and religion indicators were associated with hesitancy. The religion results are particularly informative: both religious category and the centrality of religion mattered, suggesting that the importance of religion to self-definition may provide information beyond membership in any one religious group. The sexual-orientation result illustrates the same point: membership alone did not account for the observed association, whereas high centrality was associated with both outcomes. One of the main contributions of this study is therefore a measurement one: group categorisation is more informative when identity category membership, identity centrality, and multidimensional structure are analysed separately. This extends prior work on under-vaccinated groups, including evidence that reasons for non-vaccination vary across groups, from beliefs about natural immunity and religious providence to discrimination, mobility, and access barriers (24). Crawshaw et al. (12) similarly showed that under-vaccination among migrants is shaped by access barriers, cultural acceptability, social norms, stigma, distrust, informal information sources, and beliefs about natural remedies or immunity. Membership and centrality should therefore not be treated as equivalent; disentangling the two provides additional insight into vaccination behaviour. For public health, the practical implication is that people sharing the same demographic label may differ in how strongly that identity shapes trust, perceived norms, messenger credibility, and responses to vaccine communication.

The associations observed for political affiliation, online affiliation, and lifestyle identities are consistent with literature showing that vaccine hesitancy is shaped not only by socioeconomic position or ethnicity, but also by information environments, social norms, and health worldviews (5, 24, 30, 52). In particular, online affiliations may influence exposure to vaccine narratives and trusted peer communities, while natural-health and alternative-medicine identities have previously been linked to concerns about biomedical interventions and preference for “natural” immunity (24, 52). Our findings extend this literature by defining these identities more precisely, measuring them alongside routine demographic indicators within the same analytic framework, and linking them specifically to hesitancy towards routine childhood immunisation.

A further finding is that attitudinal hesitancy did not always translate into behaviour. This aligns with prior literature showing that vaccine attitudes, intentions, and vaccination behaviour are related but distinct constructs influenced by different psychological and structural factors (5, 26, 30, 43, 54), and that, consequently, hesitant views do not necessarily translate directly into behaviour.

We used PCA descriptively to examine whether different identity dimensions tended to cluster together, rather than to classify participants into discrete identity types. This approach is consistent with social psychological perspectives emphasizing that people hold multiple social identities simultaneously and that these identities may vary in their perceived overlap, compatibility, and organization (27, 41). In particular, identity-based motivation theory suggests that health-related decisions are influenced by whether a behaviour feels congruent with salient and meaningful identities (37, 38). Similarly, work on multiple group memberships and identity compatibility shows that outcomes are shaped by broader identity configurations rather than by isolated category memberships (27). In the present study, PC1 captured a broad identity-engagement dimension, reflecting the covariation of issue-related, lifestyle, academic, community, and online affiliations. This component was associated with both attitudinal and behavioural hesitancy, suggesting that hesitancy may be patterned not only by single identities but also by broader configurations of engagement and non-engagement across domains.

Immunisation programmes could use these findings to refine segmentation. In addition to asking who belongs to a category, surveys could ask which identities are central and which affiliations co-occur. Short centrality items and self-reported affiliations with relevant community or online spaces, interpreted with local uptake data, may help detect hesitancy that demographic categories alone miss. Intervention design should then match messengers, channels, and framing to salient identities while avoiding deterministic labels and stigma (8,53). Public-health communication may also intentionally increase the salience of specific identities in particular contexts, for example, emphasising parental identity, professional responsibility, or community protection norms when promoting vaccination. This is consistent with evidence that people are more likely to act in identity-congruent ways when a given identity is salient (38, 46, 48), and with WHO guidance on Tailoring Immunization Programmes and the Behavioural and Social Drivers framework (53, 54).

Recent reviews of vaccination interventions in minority and under-vaccinated populations reach the same practical conclusion: one-size-fits-all approaches are weak, and tailored, multicomponent strategies are more plausible when they respond to local barriers, trusted institutions, and community dynamics (19, 20).

Several limitations are important. The Prolific sample was heterogeneous but non-probability-based, so prevalence estimates should not be treated as nationally representative. As an online-recruited panel sample, it may differ from the general UK parent population in education, digital engagement, age, and socioeconomic distribution. Behavioural hesitancy was self-reported and may be affected by recall bias or social desirability bias. The survey assessed attitudes toward routine childhood vaccination in general, whereas perceptions and decisions may differ across specific vaccines and diseases (9, 11). Although the questionnaire cited the measles-mumps-rubella (MMR) and diphtheria-tetanus-pertussis-polio-Hib-hepatitis B (6-in-1) vaccines as examples to anchor respondents’ thinking, responses may still reflect generalised attitudes rather than vaccine-specific considerations. We could not determine whether the reported identities or affiliations preceded vaccine hesitancy, how long respondents had held them, or whether online affiliation captured identity, exposure, or self-selection into particular information environments. Centrality was measured only when the corresponding identity-importance item was applicable or presented, and dichotomising centrality for analysis likely caused information loss. PCA provided a useful summary of covariance across indicators, but component labels are interpretive and should not be treated as authoritative. The component labels reflect the specific identity domains measured in this sample and should not be assumed to generalise to other populations or survey instruments.

Future work should test these identity-related drivers directly. Longitudinal surveys can examine whether centrality predicts changes in hesitancy over time; intervention studies can assess whether changes in salience, trust, or network exposure mediate programme effects; and qualitative work can clarify how respondents understand labels such as “natural lifestyle” or “not engaging”. Experimental work could examine whether making particular identities more salient in specific contexts, for example, parental responsibility, professional identity, or community protection norms, changes responsiveness to vaccination messages. Methodologically, the field needs brief validated centrality measures for routine surveys, transparent reporting standards for high-dimensional identity mapping, and direct comparisons of alternative approaches to modelling structured identities. The identity-landscape approach therefore reframes measurement rather than replacing established public-health stratification. It treats identity as structured and measurable, and may help public health move from describing who is under-vaccinated to understanding which identities, in which combinations, are most relevant to vaccine confidence, delay, and refusal.

## Summary Points

- Identity category membership, identity centrality, and identity structure are analytically distinct and should not be treated as interchangeable.
- Several identity membership not well described in the literature, such as identity lifestyles, online affiliations and political leanings, are strongly associated with vaccination attitudes and behaviour and may be helpful in devising new approaches to identifying undervaccinated populations.
- Greater identity centrality is often associated with increased engagement in identity-congruent behaviors.
- High-dimensional approaches such as PCA can reveal bundles of identitiesthat are not visible when variables are modelled one at a time.
- In our UK parent case example, low income, religiousness, lifestyle identities, and some online affiliations were associated with hesitancy ans in some instances bundled into multi-dimensional identities; while centrality added information beyond membership.

## Future Issues

- Future work should validate brief identity-centrality items that can be used in routine vaccination surveys without overburdening respondents.
- Longitudinal studies should test how identity membership, centrality and structure form in relation to vaccine confidence, delay, or refusal.
- Intervention studies should examine whether identity-informed messenging improves impact on vaccination attitudes and behaviour in relevant populations segmentation improves messenger choice, channel selection, and message framing.
- Ethical safeguards are needed so that identity-informed public-health measurement improves inclusion without reinforcing stigma or surveillance harm.

Down the line, to close the immunization gap, our approach could be used to developed targeted campaigns in underserved groups defined beyond tradition socio-demographic characteristics

## Data Availability

All data produced in the present study are available upon reasonable request to the authors

## Appendix 1. Survey domains reported in the case example

| Domain | Illustrative content | Why included |
| --- | --- | --- |
| Routine sociodemographic and family variables | Age-related identity, parental role, household or family-position variables, region and similar routine stratifiers. | Preserves comparability with standard public-health segmentation. |
| Socioeconomic position | Income categories, education, occupation or employment-related items, and deprivation-relevant markers. | Captures structural gradients repeatedly associated with vaccination. |
| Ethnocultural, migration and faith | Ethnicity, religion, migration background, and related community ties. | Reflects populations commonly discussed in under-vaccination research. |
| Lifestyle and health-worldview identities | Alternative-medicine identity, natural-lifestyle identity, and related self-described movements. | Captures value-based or self-chosen affiliations that are often absent from routine data. |
| Political and civic/community affiliations | Political-orientation items and broader community or civic identities were measured. | Adds norm environments that are not reducible to demographics alone. |
| Online-community affiliations | Self-identified platform communities such as YouTube, Instagram, TikTok, X/Twitter, and a no-online-affiliation option. | Captures digital affiliation without passive tracking. |
| Centrality items | Importance of selected identities on a 1-7 scale, measured separately from membership. | Separates self-definition from simple category endorsement. |
| Outcome measures | PACV score and self-reported vaccination delay/refusal. | Provides attitudinal and behavioural hesitancy outcomes. |

## Supplementary Tables: Associations between identities and vaccine hesitancy

Notes:

- For mutually exclusive categorical variables (e.g., gender, place of residence, education), risk ratios (RRs) compare each category to the most frequent category shown (reference).
- For non-mutually exclusive identity items (e.g., lifestyle identities, online affiliations), RRs compare endorsed vs not endorsed.
- Importance variables are dichotomised as High (importance ≥5) vs Low (importance <5) among respondents with non-missing importance; Low is the reference.
- To avoid sparse-cell instability, categories were combined or excluded so that both comparison groups had n >= 20.
- Adjusted models (shown only when the unadjusted p < 0.05) used modified Poisson regression with robust SE, adjusted for parent age, gender, education, household income, place of residence, number of children, and youngest-child age band. Primary outcomes were complete for all 1,000 respondents; importance analyses were restricted to respondents with non-missing or applicable importance scores for that domain.

**Supplementary Table S1.** Association between identity characteristics and attitudinal hesitancy.

| Identity domain/item | Category or indicator | n endorsed | % with outcome | Unadjusted RR | Unadjusted 95% CI | Unadjusted p | Adjusted RR | Adjusted 95% CI | Adjusted p |
| --- | --- | --- | --- | --- | --- | --- | --- | --- | --- |
| **Demographics** |  |  |  |  |  |  |  |  |  |
| Gender | Woman (reference) | 601 | 21.6 | 1 | — | — | — | — | — |
|  | Man | 394 | 20.1 | 0.93 | 0.72 - 1.19 | 0.551 | — | — | — |
| Parent age | Per 1-year increase | 1000 | 20.9 | 0.96 | 0.94 - 0.99 | <0.001 | 0.97 | 0.95 - 1.00 | 0.022 |
| Place of residence | Urban area / City (reference) | 414 | 23.4 | 1 | — | — | — | — | — |
|  | Small town | 411 | 17.0 | 0.73 | 0.55 - 0.96 | 0.023 | 0.74 | 0.57 - 0.98 | 0.033 |
|  | Rural area | 170 | 23.5 | 1.00 | 0.73 - 1.39 | 0.979 | — | — | — |
| **Family structure** |  |  |  |  |  |  |  |  |  |
| Number of children | Two children (reference) | 465 | 18.7 | 1 | — | — | — | — | — |
|  | One child | 348 | 20.1 | 1.08 | 0.81 - 1.43 | 0.615 | — | — | — |
|  | Three or more children | 187 | 27.8 | 1.49 | 1.10 - 2.00 | 0.009 | 1.33 | 0.98 - 1.79 | 0.066 |
| Youngest child age band | 5 - 9 (reference) | 433 | 18.2 | 1 | — | — | — | — | — |
|  | 3 - 4 | 232 | 21.6 | 1.18 | 0.86 - 1.62 | 0.302 | — | — | — |
|  | 1 - 2 | 239 | 24.7 | 1.35 | 1.00 - 1.82 | 0.047 | 1.18 | 0.85 - 1.63 | 0.319 |
|  | <1 | 96 | 21.9 | 1.20 | 0.78 - 1.84 | 0.405 | — | — | — |
| **Socioeconomic** |  |  |  |  |  |  |  |  |  |
| Education | Undergraduate (reference) | 451 | 20.2 | 1 | — | — | — | — | — |
|  | Secondary/vocational or less | 335 | 22.4 | 1.11 | 0.85 - 1.45 | 0.452 | — | — | — |
|  | Postgraduate | 214 | 20.1 | 1.00 | 0.72 - 1.38 | 0.98 | — | — | — |
| Household income | <£20,000 | 41 | 56.1 | 3.77 | 2.50 - 5.67 | <0.001 | 3.92 | 2.49 - 6.16 | <0.001 |
|  | £20,001 - 30,000 | 87 | 32.2 | 2.16 | 1.40 - 3.33 | <0.001 | 2.27 | 1.43 - 3.61 | <0.001 |
|  | £30,001 - 40,000 | 146 | 27.4 | 1.84 | 1.23 - 2.75 | 0.003 | 1.83 | 1.20 - 2.78 | 0.005 |
|  | £40,001 - 50,000 | 105 | 21.9 | 1.47 | 0.92 - 2.36 | 0.11 | — | — | — |
|  | £50,001 - 60,000 | 130 | 19.2 | 1.29 | 0.81 - 2.06 | 0.283 | — | — | — |
|  | £60,001 - 70,000 | 142 | 17.6 | 1.18 | 0.74 - 1.89 | 0.485 | — | — | — |
|  | £70,001 - 80,000 | 114 | 8.8 | 0.59 | 0.30 - 1.15 | 0.119 | — | — | — |
|  | >£80,000 (reference) | 235 | 14.9 | 1 | — | — | — | — | — |
| Occupation | Professional (reference) | 231 | 19.9 | 1 | — | — | — | — | — |
|  | Manager/executive | 197 | 18.8 | 0.94 | 0.64 - 1.39 | 0.768 | — | — | — |
|  | Technical/specialist | 141 | 15.6 | 0.78 | 0.49 - 1.24 | 0.302 | — | — | — |
|  | Administrative/clerical | 163 | 17.2 | 0.86 | 0.56 - 1.32 | 0.495 | — | — | — |
|  | Care/support | 42 | 40.5 | 2.03 | 1.30 - 3.18 | 0.002 | 1.44 | 0.87 - 2.37 | 0.154 |
|  | Retail/customer service | 44 | 13.6 | 0.68 | 0.31 - 1.50 | 0.346 | — | — | — |
|  | Skilled manual/craft | 30 | 40.0 | 2.01 | 1.21 - 3.34 | 0.007 | 1.82 | 1.02 - 3.27 | 0.044 |
|  | Manual/other paid work | 47 | 17.0 | 0.85 | 0.43 - 1.69 | 0.652 | — | — | — |
|  | Not in paid work | 105 | 31.4 | 1.58 | 1.08 - 2.31 | 0.02 | 1.06 | 0.70 - 1.61 | 0.771 |
| Identity |  |  |  |  |  |  |  |  |  |
| Nationality | British (reference) | 882 | 19.5 | 1 | — | — | — | — | — |
|  | Not British | 118 | 31.4 | 1.61 | 1.19 - 2.17 | 0.002 | 1.32 | 0.98 - 1.79 | 0.07 |
|  | Other nationality (selected) | 131 | 29.8 | 1.52 | 1.13 - 2.05 | 0.005 | 1.29 | 0.96 - 1.75 | 0.094 |
| Geographic/community connection | Country | 178 | 21.3 | 0.97 | 0.68 - 1.36 | 0.845 | — | — | — |
|  | Continent | 56 | 12.5 | 0.57 | 0.28 - 1.16 | 0.121 | — | — | — |
|  | World | 175 | 26.9 | 1.22 | 0.89 - 1.67 | 0.224 | — | — | — |
|  | Region | 89 | 21.3 | 0.97 | 0.62 - 1.51 | 0.88 | — | — | — |
|  | City or town | 158 | 13.9 | 0.63 | 0.41 - 0.97 | 0.038 | 0.66 | 0.43 - 1.03 | 0.066 |
|  | Local area (reference) | 344 | 22.1 | 1 | — | — | — | — | — |
| Religion or belief | Non-religious (reference) | 594 | 15.7 | 1 | — | — | — | — | — |
|  | Christian | 322 | 25.2 | 1.61 | 1.23 - 2.09 | <0.001 | 1.58 | 1.21 - 2.05 | <0.001 |
|  | Muslim | 54 | 42.6 | 2.72 | 1.90 - 3.91 | <0.001 | 1.89 | 1.28 - 2.79 | 0.001 |
|  | Other religion | 30 | 40.0 | 2.55 | 1.59 - 4.11 | <0.001 | 2.52 | 1.60 - 3.97 | <0.001 |
| Political affiliation | Left (reference) | 341 | 13.8 | 1 | — | — | — | — | — |
|  | Centrist | 314 | 21.0 | 1.53 | 1.08 - 2.15 | 0.015 | 1.41 | 1.01 - 1.97 | 0.045 |
|  | Right | 164 | 28.0 | 2.04 | 1.42 - 2.92 | <0.001 | 1.89 | 1.33 - 2.69 | <0.001 |
|  | No affiliation | 168 | 29.2 | 2.12 | 1.48 - 3.02 | <0.001 | 1.88 | 1.30 - 2.72 | <0.001 |
| Ethnicity | White (reference) | 815 | 17.2 | 1 | — | — | — | — | — |
|  | Black | 71 | 45.1 | 2.62 | 1.95 - 3.53 | <0.001 | 2.30 | 1.59 - 3.31 | <0.001 |
|  | Asian | 70 | 32.9 | 1.91 | 1.32 - 2.76 | <0.001 | 1.64 | 1.12 - 2.42 | 0.012 |
|  | Mixed/Other | 44 | 31.8 | 1.85 | 1.17 - 2.93 | 0.008 | 1.82 | 1.14 - 2.89 | 0.012 |
| Sexual orientation | Straight/heterosexual (reference) | 922 | 21.4 | 1 | — | — | — | — | — |
|  | Sexual minority | 78 | 15.4 | 0.72 | 0.42 - 1.23 | 0.229 | — | — | — |
| Affiliations |  |  |  |  |  |  |  |  |  |
| Lifestyle affiliation | Vegan or vegetarian | 89 | 21.3 | 1.02 | 0.67 - 1.56 | 0.913 | — | — | — |
|  | Natural lifestyle | 118 | 43.2 | 2.41 | 1.88 - 3.10 | <0.001 | 2.33 | 1.80 - 3.03 | <0.001 |
|  | Preference for alternative medicine | 63 | 60.3 | 3.31 | 2.60 - 4.21 | <0.001 | 3.17 | 2.40 - 4.18 | <0.001 |
|  | Minimalist lifestyle | 125 | 32.8 | 1.71 | 1.28 - 2.27 | <0.001 | 1.52 | 1.15 - 2.01 | 0.003 |
|  | Environmentally/climate-conscious lifestyle | 205 | 26.3 | 1.35 | 1.03 - 1.77 | 0.028 | 1.30 | 0.99 - 1.70 | 0.061 |
|  | None of the above | 627 | 16.1 | 0.56 | 0.44 - 0.71 | <0.001 | 0.57 | 0.45 - 0.73 | <0.001 |
| Identity: Online communities |  |  |  |  |  |  |  |  |  |
|  | Gamer affiliation | 181 | 23.2 | 1.14 | 0.85 - 1.53 | 0.395 | — | — | — |
|  | TikTok affiliation | 79 | 35.4 | 1.80 | 1.30 - 2.50 | <0.001 | 1.38 | 0.99 - 1.93 | 0.06 |
|  | YouTube affiliation | 69 | 40.6 | 2.09 | 1.52 - 2.86 | <0.001 | 1.73 | 1.23 - 2.45 | 0.002 |
|  | Reddit affiliation | 114 | 16.7 | 0.78 | 0.51 - 1.19 | 0.25 | — | — | — |
|  | Music fandom | 38 | 18.4 | 0.88 | 0.44 - 1.73 | 0.706 | — | — | — |
|  | Crypto community | 24 | 37.5 | 1.83 | 1.08 - 3.11 | 0.026 | 1.81 | 1.00 - 3.25 | 0.048 |
|  | Parenting/health forums | 84 | 23.8 | 1.15 | 0.77 - 1.73 | 0.486 | — | — | — |
|  | Telegram/activist/other online affiliation (combined) | 37 | 29.7 | 1.45 | 0.87 - 2.41 | 0.157 | — | — | — |
|  | Instagram affiliation | 247 | 29.6 | 1.64 | 1.28 - 2.09 | <0.001 | 1.51 | 1.18 - 1.94 | <0.001 |
|  | Facebook affiliation | 205 | 25.4 | 1.28 | 0.98 - 1.69 | 0.073 | — | — | — |
|  | X/Twitter affiliation | 73 | 38.4 | 1.96 | 1.43 - 2.70 | <0.001 | 1.95 | 1.38 - 2.75 | <0.001 |
|  | No online affiliation | 477 | 16.4 | 0.65 | 0.51 - 0.84 | <0.001 | 0.69 | 0.54 - 0.88 | 0.003 |
| Ideology/cause affiliation | Feminism/gender equality | 318 | 17.3 | 0.77 | 0.58 - 1.01 | 0.06 | — | — | — |
|  | Racial equality | 332 | 20.5 | 0.97 | 0.75 - 1.26 | 0.819 | — | — | — |
|  | Religious tolerance/interfaith | 185 | 19.5 | 0.92 | 0.66 - 1.26 | 0.596 | — | — | — |
|  | Climate change mitigation | 298 | 17.8 | 0.80 | 0.60 - 1.06 | 0.12 | — | — | — |
|  | Human rights | 400 | 24.8 | 1.35 | 1.06 - 1.72 | 0.014 | 1.21 | 0.95 - 1.55 | 0.12 |
|  | Animal rights | 291 | 21.0 | 1.00 | 0.77 - 1.31 | 0.975 | — | — | — |
|  | No issue engagement | 429 | 19.3 | 0.88 | 0.68 - 1.12 | 0.297 | — | — | — |
| Academic affiliation | University student | 68 | 26.5 | 1.29 | 0.85 - 1.96 | 0.228 | — | — | — |
|  | University alumni society member | 119 | 27.7 | 1.39 | 1.01 - 1.91 | 0.044 | 1.39 | 0.98 - 1.97 | 0.061 |
|  | University staff and/or academic research group | 43 | 18.6 | 0.89 | 0.47 - 1.68 | 0.709 | — | — | — |
|  | No academic affiliation | 783 | 19.4 | 0.74 | 0.57 - 0.96 | 0.025 | 0.74 | 0.55 - 0.99 | 0.04 |
| Community affiliation | Volunteer | 181 | 21.0 | 1.01 | 0.74 - 1.37 | 0.972 | — | — | — |
|  | Union member | 67 | 22.4 | 1.08 | 0.68 - 1.71 | 0.754 | — | — | — |
|  | Community leader/activist/other role (combined) | 25 | 44.0 | 2.17 | 1.37 - 3.43 | <0.001 | 1.64 | 1.06 - 2.54 | 0.026 |
|  | No community role | 749 | 20.3 | 0.89 | 0.68 - 1.17 | 0.412 | — | — | — |
| Cultural affiliation | Musical subculture | 208 | 17.3 | 0.79 | 0.57 - 1.10 | 0.16 | — | — | — |
|  | Sports fan/team supporter | 390 | 19.7 | 0.91 | 0.71 - 1.17 | 0.473 | — | — | — |
|  | Artistic/creative community | 153 | 24.8 | 1.23 | 0.91 - 1.67 | 0.185 | — | — | — |
|  | Other interest/cultural (specified) | 32 | 15.6 | 0.74 | 0.33 - 1.67 | 0.471 | — | — | — |
|  | No cultural/interest group | 419 | 21.2 | 1.03 | 0.81 - 1.31 | 0.822 | — | — | — |

**Supplementary Table S2.** Association between identity importance and attitudinal hesitancy.

| Identity domain/item | Category or indicator | n endorsed | % with outcome | Unadjusted RR | Unadjusted 95% CI | Unadjusted p | Adjusted RR | Adjusted 95% CI | Adjusted p |
| --- | --- | --- | --- | --- | --- | --- | --- | --- | --- |
| Gender importance | High (importance $\geq 5$) | 887 | 20.7 | 0.94 | 0.65 - 1.36 | 0.732 | — | — | — |
| | Low (importance $< 5$) (reference) | 113 | 22.1 | 1.0 | — | — | — | — | — |
| Age importance | High (importance $\geq 5$) | 479 | 24.8 | 1.44 | 1.13 - 1.84 | 0.004 | 1.67 | 1.07 - 2.61 | 0.011 |
| | Low (importance $< 5$) (reference) | 521 | 17.3 | 1.0 | — | — | — | — | — |
| Place of residence importance | High (importance $\geq 5$) | 540 | 22.2 | 1.15 | 0.90 - 1.47 | 0.267 | — | — | — |
| | Low (importance $< 5$) (reference) | 460 | 19.3 | 1.0 | — | — | — | — | — |
| Education importance | High (importance $\geq 5$) | 598 | 20.9 | 1.00 | 0.78 - 1.28 | 0.998 | — | — | — |
| | Low (importance $< 5$) (reference) | 402 | 20.9 | 1.0 | — | — | — | — | — |
| Income importance | High (importance $\geq 5$) | 523 | 22.8 | 1.21 | 0.94 - 1.54 | 0.133 | — | — | — |
| | Low (importance $< 5$) (reference) | 477 | 18.9 | 1.0 | — | — | — | — | — |
| Occupation importance | High (importance $\geq 5$) | 702 | 20.2 | 0.90 | 0.70 - 1.16 | 0.42 | — | — | — |
|  | Low (importance <5) (reference) | 298 | 22.5 | 1.0 | — | — | — | — | — |
| Nationality importance | High (importance ≥5) | 712 | 21.1 | 1.03 | 0.79 - 1.34 | 0.838 | — | — | — |
|  | Low (importance <5) (reference) | 288 | 20.5 | 1.0 | — | — | — | — | — |
| Immigration history importance | High (importance ≥5) | 72 | 36.1 | 1.30 | 0.71 - 2.39 | 0.399 | — | — | — |
|  | Low (importance <5) (reference) | 36 | 27.8 | 1.0 | — | — | — | — | — |
| Overall Geographic connection importance | High (importance ≥5) | 701 | 22.3 | 1.26 | 0.95 - 1.66 | 0.112 | — | — | — |
|  | Low (importance <5) (reference) | 299 | 17.7 | 1.0 | — | — | — | — | — |
| Importance of connection with local area | High (importance ≥5) | 247 | 21.9 | 0.96 | 0.62 - 1.49 | 0.869 | — | — | — |
|  | Low (importance <5) (reference) | 97 | 22.7 | 1.0 | — | — | — | — | — |
| Importance of connection with city/town | High (importance ≥5) | 98 | 14.3 | 1.07 | 0.48 - 2.40 | 0.867 | — | — | — |
|  | Low (importance <5) (reference) | 60 | 13.3 | 1.0 | — | — | — | — | — |
| Importance of connection with region | High (importance ≥5) | 65 | 24.6 | 1.97 | 0.63 - 6.16 | 0.244 | — | — | — |
|  | Low (importance <5) (reference) | 24 | 12.5 | 1.0 | — | — | — | — | — |
| Importance of connection with country | High (importance ≥5) | 119 | 25.2 | 1.86 | 0.91 - 3.80 | 0.089 | — | — | — |
|  | Low (importance <5) (reference) | 59 | 13.6 | 1.0 | — | — | — | — | — |
| Importance of connection with continent | High (importance ≥5) | 36 | 19.4 | 8.51 | 0.51 - 141.72 | 0.136 | — | — | — |
|  | Low (importance <5) (reference) | 20 | 0.0 | 1.0 | — | — | — | — | — |
| Importance of connection with world | High (importance ≥5) | 136 | 25.7 | 0.84 | 0.48 - 1.45 | 0.525 | — | — | — |
|  | Low (importance <5) (reference) | 39 | 30.8 | 1.0 | — | — | — | — | — |
| Religion importance | High (importance ≥5) | 260 | 36.5 | 2.54 | 1.66 - 3.89 | <0.001 | 2.20 | 1.43 - 3.37 | <0.001 |
|  | Low (importance <5) (reference) | 146 | 14.4 | 1.0 | — | — | — | — | — |
| Political affiliation importance | High (importance ≥5) | 416 | 20.9 | 1.19 | 0.90 - 1.58 | 0.219 | — | — | — |
|  | Low (importance <5) (reference) | 416 | 17.5 | 1.0 | — | — | — | — | — |
| Importance of not being politically affiliated | High (importance ≥5) | 34 | 35.3 | 1.28 | 0.75 - 2.17 | 0.365 | — | — | — |
|  | Low (importance <5) (reference) | 134 | 27.6 | 1.0 | — | — | — | — | — |
| Ethnicity importance | High (importance ≥5) | 697 | 22.5 | 1.31 | 0.99 - 1.74 | 0.06 | — | — | — |
|  | Low (importance <5) (reference) | 303 | 17.2 | 1.0 | — | — | — | — | — |
| **Lifestyle importance (2nd selected)** |  |  |  |  |  |  |  |  |  |
| Lifestyle affiliation importance (2nd selected) | High (importance ≥5) | 173 | 20.8 | 0.98 | 0.53 - 1.82 | 0.944 | — | — | — |
| Lifestyle importance (2nd selected) | Low (importance <5) (reference) | 47 | 21.3 | 1.0 | — | — | — | — | — |
| Sexual orientation importance | High (importance ≥5) | 685 | 24.1 | 1.72 | 1.27 - 2.34 | <0.001 | 1.56 | 1.15 - 2.10 | 0.004 |
|  | Low (importance <5) (reference) | 315 | 14.0 | 1.0 | — | — | — | — | — |
| Cultural group affiliation importance (1st selected) | High (importance ≥5) | 323 | 20.7 | 0.87 | 0.57 - 1.34 | 0.523 | — | — | — |
| Cultural group importance (1st selected) | Low (importance <5) (reference) | 88 | 23.9 | 1.0 | — | — | — | — | — |
| Online affiliation importance | High (importance ≥5) | 284 | 29.9 | 1.56 | 1.14 - 2.13 | 0.006 | 1.38 | 1.00 - 1.90 | 0.048 |
|  | Low (importance <5) (reference) | 239 | 19.2 | 1.0 | — | — | — | — | — |
| Ideology participation importance | High (importance $\geq 5$) | 463 | 22.5 | 1.10 | 0.73 - 1.66 | 0.64 | — | — | — |
| | Low (importance $< 5$) (reference) | 108 | 20.4 | 1.0 | — | — | — | — | — |
| Not engaging in lifestyle or ideologies importance | High (importance $\geq 5$) | 64 | 43.8 | 2.90 | 2.01 - 4.20 | $< 0.001$ | 2.61 | 1.81 - 3.78 | $< 0.001$ |
| | Low (importance $< 5$) (reference) | 365 | 15.1 | 1.0 | — | — | — | — | — |
| Academic affiliation importance | High (importance $\geq 5$) | 127 | 31.5 | 1.67 | 1.01 - 2.75 | 0.045 | 1.43 | 0.88 - 2.33 | 0.149 |
| | Low (importance $< 5$) (reference) | 90 | 18.9 | 1.0 | — | — | — | — | — |
| Community role importance | High (importance $\geq 5$) | 186 | 24.7 | 1.46 | 0.81 - 2.65 | 0.211 | — | — | — |
| | Low (importance $< 5$) (reference) | 65 | 16.9 | 1.0 | — | — | — | — | — |

**Supplementary Table S3.** Association between identity characteristics and behavioural hesitancy.

| Identity domain/item | Category or indicator | n endorsed | % with outcome | Unadjusted RR | Unadjusted 95% CI | Unadjusted p | Adjusted RR | Adjusted 95% CI | Adjusted p |
| --- | --- | --- | --- | --- | --- | --- | --- | --- | --- |
| **Demographics** |  |  |  |  |  |  |  |  |  |
| Gender | Woman (reference) | 601 | 9.2 | 1 | — | — | — | — | — |
|  | Man | 394 | 6.3 | 0.69 | 0.44 - 1.09 | 0.115 | — | — | — |
| Parent age | Per 1-year increase | 1000 | 8.0 | 0.96 | 0.93 - 0.99 | 0.017 | 0.96 | 0.93 - 1.01 | 0.094 |
| Place of residence | Urban area / City (reference) | 414 | 10.4 | 1 | — | — | — | — | — |
|  | Small town | 411 | 5.8 | 0.56 | 0.35 - 0.91 | 0.019 | 0.56 | 0.34 - 0.92 | 0.021 |
|  | Rural area | 170 | 7.6 | 0.74 | 0.41 - 1.33 | 0.312 | — | — | — |
| **Family structure** |  |  |  |  |  |  |  |  |  |
| Number of children | Two children (reference) | 465 | 7.7 | 1 | — | — | — | — | — |
|  | One child | 348 | 6.6 | 0.85 | 0.52 - 1.41 | 0.539 | — | — | — |
|  | Three or more children | 187 | 11.2 | 1.45 | 0.87 - 2.42 | 0.153 | — | — | — |
| Youngest child age band | 5 - 9 (reference) | 433 | 6.0 | 1 | — | — | — | — | — |
|  | 3 - 4 | 232 | 9.1 | 1.51 | 0.87 - 2.62 | 0.145 | — | — | — |
|  | 1 - 2 | 239 | 10.9 | 1.81 | 1.08 - 3.05 | 0.025 | 1.44 | 0.81 - 2.57 | 0.214 |
| | $< 1$ | 96 | 7.3 | 1.21 | 0.54 - 2.72 | 0.636 | — | — | — |
| **Socioeconomic** |  |  |  |  |  |  |  |  |  |
| Education | Undergraduate (reference) | 451 | 8.9 | 1 | — | — | — | — | — |
|  | Secondary/vocational or less | 335 | 6.3 | 0.71 | 0.42 - 1.18 | 0.181 | — | — | — |
|  | Postgraduate | 214 | 8.9 | 1.00 | 0.59 - 1.69 | 0.997 | — | — | — |
| Income category | $< £20,000$ | 41 | 19.5 | 3.28 | 1.47 - 7.31 | 0.004 | 3.38 | 1.43 - 8.03 | 0.006 |
|  | £20,001 - 30,000 | 87 | 10.3 | 1.74 | 0.78 - 3.87 | 0.177 | — | — | — |
|  | £30,001 - 40,000 | 146 | 7.5 | 1.26 | 0.59 - 2.71 | 0.546 | — | — | — |
|  | £40,001 - 50,000 | 105 | 8.6 | 1.44 | 0.64 - 3.22 | 0.376 | — | — | — |
|  | £50,001 - 60,000 | 130 | 8.5 | 1.42 | 0.66 - 3.04 | 0.366 | — | — | — |
|  | £60,001 - 70,000 | 142 | 8.5 | 1.42 | 0.68 - 2.98 | 0.356 | — | — | — |
|  | £70,001 - 80,000 | 114 | 5.3 | 0.88 | 0.35 - 2.24 | 0.794 | — | — | — |
| | $> £80,000$ (reference) | 235 | 6.0 | 1 | — | — | — | — | — |
| Occupation | Professional (reference) | 231 | 10.0 | 1 | — | — | — | — | — |
|  | Manager/executive | 197 | 6.6 | 0.66 | 0.34 - 1.27 | 0.217 | — | — | — |
|  | Technical/specialist | 141 | 6.4 | 0.64 | 0.31 - 1.35 | 0.24 | — | — | — |
|  | Administrative/clerical | 163 | 5.5 | 0.55 | 0.26 - 1.17 | 0.12 | — | — | — |
|  | Care/support | 42 | 16.7 | 1.67 | 0.77 - 3.65 | 0.195 | — | — | — |
|  | Retail/customer service | 44 | 2.3 | 0.23 | 0.03 - 1.65 | 0.143 | — | — | — |
|  | Skilled manual/craft | 30 | 3.3 | 0.33 | 0.05 - 2.39 | 0.275 | — | — | — |
|  | Manual/other paid work | 47 | 6.4 | 0.64 | 0.20 - 2.05 | 0.453 | — | — | — |
|  | Not in paid work | 105 | 13.3 | 1.34 | 0.72 - 2.50 | 0.358 | — | — | — |
| **Identity** |  |  |  |  |  |  |  |  |  |
| Nationality | British (reference) | 882 | 7.8 | 1 | — | — | — | — | — |
|  | Not British | 118 | 9.3 | 1.19 | 0.65 - 2.19 | 0.571 | — | — | — |
|  | Other nationality (selected) | 131 | 9.2 | 1.17 | 0.65 - 2.10 | 0.598 | — | — | — |
| Geographic/community connection | Country | 178 | 7.3 | 0.90 | 0.48 - 1.69 | 0.737 | — | — | — |
|  | Continent | 56 | 8.9 | 1.10 | 0.44 - 2.72 | 0.842 | — | — | — |
|  | World | 175 | 10.9 | 1.33 | 0.77 - 2.32 | 0.308 | — | — | — |
|  | Region | 89 | 7.9 | 0.97 | 0.44 - 2.14 | 0.933 | — | — | — |
|  | City or town | 158 | 5.1 | 0.62 | 0.29 - 1.33 | 0.223 | — | — | — |
|  | Local area (reference) | 344 | 8.1 | 1 | — | — | — | — | — |
| Religion or belief | Non-religious (reference) | 594 | 5.6 | 1 | — | — | — | — | — |
|  | Christian | 322 | 11.8 | 2.12 | 1.36 - 3.32 | <0.001 | 1.96 | 1.25 - 3.07 | 0.003 |
|  | Muslim | 54 | 11.1 | 2.00 | 0.88 - 4.56 | 0.099 | — | — | — |
|  | Other religion | 30 | 10.0 | 1.80 | 0.59 - 5.54 | 0.305 | — | — | — |
| Political affiliation | Left (reference) | 341 | 7.6 | 1 | — | — | — | — | — |
|  | Centrist | 314 | 8.0 | 1.04 | 0.62 - 1.77 | 0.872 | — | — | — |
|  | Right | 164 | 9.1 | 1.20 | 0.65 - 2.20 | 0.557 | — | — | — |
|  | No affiliation | 168 | 8.3 | 1.09 | 0.59 - 2.04 | 0.78 | — | — | — |
| Ethnicity | White (reference) | 815 | 6.4 | 1 | — | — | — | — | — |
|  | Black | 71 | 16.9 | 2.65 | 1.48 - 4.73 | <0.001 | 1.91 | 0.97 - 3.73 | 0.06 |
|  | Asian | 70 | 14.3 | 2.24 | 1.19 - 4.21 | 0.012 | 1.61 | 0.85 - 3.04 | 0.143 |
|  | Mixed/Other | 44 | 13.6 | 2.14 | 0.97 - 4.70 | 0.059 | — | — | — |
| Sexual orientation | Straight/heterosexual (reference) | 922 | 8.0 | 1 | — | — | — | — | — |
|  | Sexual minority | 78 | 7.7 | 0.96 | 0.43 - 2.13 | 0.917 | — | — | — |
| **Affiliations** |  |  |  |  |  |  |  |  |  |
| Lifestyle affiliation | Vegan or vegetarian | 89 | 7.9 | 0.98 | 0.47 - 2.07 | 0.961 | — | — | — |
|  | Natural lifestyle | 118 | 20.3 | 3.20 | 2.07 - 4.96 | <0.001 | 2.73 | 1.68 - 4.45 | <0.001 |
|  | Preference for alternative medicine | 63 | 28.6 | 4.32 | 2.73 - 6.83 | <0.001 | 3.98 | 2.35 - 6.75 | <0.001 |
|  | Minimalist lifestyle | 125 | 14.4 | 2.03 | 1.24 - 3.32 | 0.005 | 1.65 | 0.98 - 2.79 | 0.06 |
|  | Environmentally/climate-conscious lifestyle | 205 | 11.2 | 1.56 | 0.99 - 2.48 | 0.056 | — | — | — |
|  | None of the above | 627 | 5.6 | 0.46 | 0.30 - 0.71 | <0.001 | 0.53 | 0.34 - 0.82 | 0.005 |
| **Identity: Online communities** |  |  |  |  |  |  |  |  |  |
|  | Gamer affiliation | 181 | 8.8 | 1.13 | 0.67 - 1.91 | 0.644 | — | — | — |
|  | TikTok affiliation | 79 | 19.0 | 2.69 | 1.61 - 4.49 | <0.001 | 2.24 | 1.33 - 3.77 | 0.002 |
|  | YouTube affiliation | 69 | 15.9 | 2.15 | 1.20 - 3.87 | 0.011 | 2.02 | 1.14 - 3.58 | 0.017 |
|  | Reddit affiliation | 114 | 8.8 | 1.11 | 0.59 - 2.09 | 0.746 | — | — | — |
|  | Music fandom | 38 | 7.9 | 0.99 | 0.33 - 2.98 | 0.981 | — | — | — |
|  | Crypto community | 24 | 8.3 | 1.04 | 0.27 - 4.00 | 0.951 | — | — | — |
|  | Parenting/health forums | 84 | 7.1 | 0.88 | 0.40 - 1.97 | 0.763 | — | — | — |
|  | Telegram/activist/other online affiliation (combined) | 37 | 13.5 | 1.74 | 0.75 - 4.03 | 0.2 | — | — | — |
|  | Instagram affiliation | 247 | 11.7 | 1.73 | 1.12 - 2.67 | 0.013 | 1.57 | 1.03 - 2.39 | 0.034 |
|  | Facebook affiliation | 205 | 11.2 | 1.56 | 0.99 - 2.48 | 0.056 | — | — | — |
|  | X/Twitter affiliation | 73 | 15.1 | 2.02 | 1.12 - 3.65 | 0.019 | 2.19 | 1.17 - 4.10 | 0.014 |
|  | No online affiliation | 477 | 6.7 | 0.73 | 0.48 - 1.12 | 0.153 | — | — | — |
| **Ideology / cause participation** | Feminism/gender equality | 318 | 9.4 | 1.29 | 0.83 - 1.98 | 0.253 | — | — | — |
|  | Racial equality | 332 | 10.2 | 1.49 | 0.97 - 2.27 | 0.066 | — | — | — |
|  | Religious tolerance/interfaith | 185 | 9.7 | 1.28 | 0.78 - 2.11 | 0.335 | — | — | — |
|  | Climate change mitigation | 298 | 7.4 | 0.89 | 0.56 - 1.43 | 0.64 | — | — | — |
|  | Human rights | 400 | 10.5 | 1.66 | 1.09 - 2.52 | 0.018 | 1.38 | 0.88 - 2.18 | 0.165 |
|  | Animal rights | 291 | 7.2 | 0.87 | 0.54 - 1.40 | 0.56 | — | — | — |
|  | No issue engagement | 429 | 7.2 | 0.84 | 0.55 - 1.30 | 0.435 | — | — | — |
| Academic affiliation | University student | 68 | 14.7 | 1.96 | 1.06 - 3.62 | 0.032 | 1.65 | 0.86 - 3.17 | 0.13 |
|  | University alumni society member | 119 | 13.4 | 1.85 | 1.11 - 3.09 | 0.019 | 1.63 | 0.95 - 2.81 | 0.078 |
|  | University staff and/or academic research group | 43 | 4.7 | 0.57 | 0.15 - 2.25 | 0.422 | — | — | — |
|  | No academic affiliation | 783 | 6.9 | 0.58 | 0.37 - 0.90 | 0.015 | 0.66 | 0.40 - 1.08 | 0.097 |
| Community affiliation | Volunteer | 181 | 7.2 | 0.88 | 0.50 - 1.56 | 0.655 | — | — | — |
|  | Union member | 67 | 14.9 | 1.99 | 1.08 - 3.68 | 0.028 | 2.28 | 1.20 - 4.34 | 0.012 |
|  | Community leader/activist/other role (combined) | 25 | 24.0 | 3.16 | 1.52 - 6.57 | 0.002 | 2.28 | 1.06 - 4.88 | 0.034 |
|  | No community role | 749 | 7.3 | 0.74 | 0.47 - 1.16 | 0.185 | — | — | — |
| Cultural affiliation | Musical subculture | 208 | 7.7 | 0.95 | 0.56 - 1.61 | 0.854 | — | — | — |
|  | Sports fan/team supporter | 390 | 8.5 | 1.10 | 0.72 - 1.68 | 0.667 | — | — | — |
|  | Artistic/creative community | 153 | 8.5 | 1.07 | 0.61 - 1.90 | 0.805 | — | — | — |
|  | Other interest/cultural (specified) | 32 | 9.4 | 1.18 | 0.39 - 3.53 | 0.769 | — | — | — |
|  | No cultural/interest group | 419 | 6.7 | 0.75 | 0.48 - 1.16 | 0.195 | — | — | — |

**Supplementary Table S4.** Association between identity importance and behavioural hesitancy.

| Identity domain/item | Category or indicator | n endorsed | % with outcome | Unadjusted RR | Unadjusted 95% CI | Unadjusted p | Adjusted RR | Adjusted 95% CI | Adjusted p |
| --- | --- | --- | --- | --- | --- | --- | --- | --- | --- |
| Gender importance | High (importance $\geq 5$) | 887 | 7.9 | 0.89 | 0.47 - 1.68 | 0.723 | — | — | — |
| | Low (importance $< 5$) (reference) | 113 | 8.8 | 1.0 | — | — | — | — | — |
| Age importance | High (importance $\geq 5$) | 479 | 10.6 | 1.91 | 1.23 - 2.97 | 0.004 | 1.67 | 1.07 - 2.61 | 0.025 |
| | Low (importance $< 5$) (reference) | 521 | 5.6 | 1.0 | — | — | — | — | — |
| Place of residence importance | High (importance $\geq 5$) | 540 | 8.9 | 1.28 | 0.83 - 1.96 | 0.264 | — | — | — |
| | Low (importance $< 5$) (reference) | 460 | 7.0 | 1.0 | — | — | — | — | — |
| Education importance | High (importance $\geq 5$) | 598 | 8.7 | 1.25 | 0.80 - 1.94 | 0.325 | — | — | — |
| | Low (importance $< 5$) (reference) | 402 | 7.0 | 1.0 | — | — | — | — | — |
| Income importance | High (importance $\geq 5$) | 523 | 8.2 | 1.06 | 0.70 - 1.62 | 0.787 | — | — | — |
| | Low (importance $< 5$) (reference) | 477 | 7.8 | 1.0 | — | — | — | — | — |
| Occupation importance | High (importance $\geq 5$) | 702 | 8.4 | 1.19 | 0.74 - 1.93 | 0.471 | — | — | — |
| | Low (importance $< 5$) (reference) | 298 | 7.0 | 1.0 | — | — | — | — | — |
| Nationality importance | High (importance $\geq 5$) | 712 | 8.4 | 1.21 | 0.75 - 1.98 | 0.436 | — | — | — |
| | Low (importance $< 5$) (reference) | 288 | 6.9 | 1.0 | — | — | — | — | — |
| Immigration history importance | High (importance $\geq 5$) | 72 | 11.1 | 1.00 | 0.32 - 3.10 | 1 | — | — | — |
| | Low (importance $< 5$) (reference) | 36 | 11.1 | 1.0 | — | — | — | — | — |
| Overall Geographic connection importance | High (importance $\geq 5$) | 701 | 8.6 | 1.28 | 0.79 - 2.08 | 0.322 | — | — | — |
| | Low (importance $< 5$) (reference) | 299 | 6.7 | 1.0 | — | — | — | — | — |
| Importance of connection with local area | High (importance $\geq 5$) | 247 | 8.1 | 0.98 | 0.45 - 2.15 | 0.963 | — | — | — |
| | Low (importance $< 5$) (reference) | 97 | 8.2 | 1.0 | — | — | — | — | — |
| Importance of connection with city/town | High (importance $\geq 5$) | 98 | 5.1 | 1.02 | 0.25 - 4.12 | 0.977 | — | — | — |
| | Low (importance $< 5$) (reference) | 60 | 5.0 | 1.0 | — | — | — | — | — |
| Importance of connection with region | High (importance $\geq 5$) | 65 | 7.7 | 0.92 | 0.19 - 4.44 | 0.92 | — | — | — |
| | Low (importance $< 5$) (reference) | 24 | 8.3 | 1.0 | — | — | — | — | — |
| Importance of connection with country | High (importance $\geq 5$) | 119 | 9.2 | 2.73 | 0.62 - 11.91 | 0.182 | — | — | — |
| | Low (importance $< 5$) (reference) | 59 | 3.4 | 1.0 | — | — | — | — | — |
| Importance of connection with continent | High (importance $\geq 5$) | 36 | 13.9 | 6.24 | 0.36 - 107.41 | 0.207 | — | — | — |
| | Low (importance $< 5$) (reference) | 20 | 0.0 | 1.0 | — | — | — | — | — |
| Importance of connection with world | High (importance $\geq 5$) | 136 | 10.3 | 0.80 | 0.31 - 2.09 | 0.653 | — | — | — |
| | Low (importance $< 5$) (reference) | 39 | 12.8 | 1.0 | — | — | — | — | — |
| Religion importance | High (importance $\geq 5$) | 260 | 14.6 | 2.37 | 1.18 - 4.76 | 0.015 | 1.89 | 0.93 - 3.84 | 0.078 |
| | Low (importance $< 5$) (reference) | 146 | 6.2 | 1.0 | — | — | — | — | — |
| Political affiliation importance | High (importance $\geq 5$) | 416 | 9.9 | 1.64 | 1.02 - 2.65 | 0.043 | 1.70 | 1.04 - 2.77 | 0.034 |
| | Low (importance $< 5$) (reference) | 416 | 6.0 | 1.0 | — | — | — | — | — |
| Importance of not being politically affiliated | High (importance $\geq 5$) | 34 | 17.6 | 2.96 | 1.10 - 7.95 | 0.032 | 4.25 | 1.30 - 13.88 | 0.017 |
| | Low (importance $< 5$) (reference) | 134 | 6.0 | 1.0 | — | — | — | — | — |
| Ethnicity importance | High (importance $\geq 5$) | 697 | 8.8 | 1.40 | 0.85 - 2.29 | 0.189 | — | — | — |
| | Low (importance $< 5$) (reference) | 303 | 6.3 | 1.0 | — | — | — | — | — |
| **Lifestyle importance (2nd selected)** |  |  |  |  |  |  |  |  |  |
| Lifestyle importance (2nd selected) | High (importance $\geq 5$) | 173 | 5.8 | 0.54 | 0.20 - 1.51 | 0.243 | — | — | — |
| | Low (importance $< 5$) (reference) | 47 | 10.6 | 1.0 | — | — | — | — | — |
| Sexual orientation importance | High (importance $\geq 5$) | 685 | 9.6 | 2.17 | 1.24 - 3.80 | 0.007 | 2.03 | 1.15 - 3.60 | 0.015 |
| | Low (importance $< 5$) (reference) | 315 | 4.4 | 1.0 | — | — | — | — | — |
| Cultural group importance (1st selected) | High (importance $\geq 5$) | 323 | 9.9 | 1.09 | 0.52 - 2.28 | 0.819 | — | — | — |
| | Low (importance $< 5$) (reference) | 88 | 9.1 | 1.0 | — | — | — | — | — |
| Online affiliation importance | High (importance $\geq 5$) | 284 | 10.2 | 1.28 | 0.74 - 2.23 | 0.374 | — | — | — |
| | Low (importance $< 5$) (reference) | 239 | 7.9 | 1.0 | — | — | — | — | — |
| Ideology participation importance | High (importance $\geq 5$) | 463 | 8.2 | 0.81 | 0.43 - 1.52 | 0.507 | — | — | — |
| | Low (importance $< 5$) (reference) | 108 | 10.2 | 1.0 | — | — | — | — | — |
| Not engaging in lifestyle or ideologies importance | High (importance $\geq 5$) | 64 | 18.8 | 3.60 | 1.84 - 7.05 | <0.001 | 3.18 | 1.46 - 6.92 | 0.004 |
| | Low (importance $< 5$) (reference) | 365 | 5.2 | 1.0 | — | — | — | — | — |
| Academic affiliation importance | High (importance $\geq 5$) | 127 | 15.0 | 1.92 | 0.84 - 4.38 | 0.119 | — | — | — |
| | Low (importance $< 5$) (reference) | 90 | 7.8 | 1.0 | — | — | — | — | — |
| Community role importance | High (importance $\geq 5$) | 186 | 11.3 | 1.83 | 0.65 - 5.15 | 0.249 | — | — | — |
| | Low (importance $< 5$) (reference) | 65 | 6.2 | 1.0 | — | — | — | — | — |

**Supplementary Table S5.** PCA summary for the identity-only feature matrix.

| Component | Variance explained | Dominant loading pattern | Adjusted association with outcomes (aRR per 1 SD, 95% CI) |
| --- | --- | --- | --- |
| PC1 | 9.62% (43.77% by PC10; 54.61% by PC15) | Pos: human rights; climate mitigation; racial equality. Neg: no issue engagement; no lifestyle identity; White ethnicity. | Attitudinal 1.27 (1.14-1.42); Behavioural 1.35 (1.13-1.61). |
| PC2 | 6.06% | Positive: man; sports fan; X/Twitter. Negative: woman; feminism/gender equality; no online affiliation. | Attitudinal 0.80 (0.70-0.91); Behavioural 0.88 (0.68-1.13). |

| Component | Variance explained | Dominant loading pattern | Adjusted association with outcomes<br>(aRR per 1 SD, 95% CI) |
| --- | --- | --- | --- |
| PC3 | 5.98% | Positive: non-British nationality;<br>Black ethnicity; Muslim identity.<br>Negative: British nationality; White<br>ethnicity; non-religious identity. | Attitudinal 0.97 (0.84-1.11);<br>Behavioural 1.13 (0.89-1.43). |

Note: PCA used 65 binary identity indicators only; age, socioeconomic variables, family-structure variables, identity-importance variables, and outcomes were excluded from the feature matrix. Mutually exclusive categorical variables were one-hot encoded, checkbox variables were retained as binary indicators, and all features were standardised before PCA. Loading patterns list selected dominant positive and negative loadings rather than the full loading matrix. Outcome associations are adjusted risk ratios per 1 SD increase in component score from modified Poisson models with robust standard errors, adjusted for parent age, gender, place of residence, number of children, youngest child age band, education, and household income.

## Notes

### Competing Interest Statement

The authors have declared no competing interest.

### Author Declarations

Bar-Ilan University Faculty of Medicine Ethics committee

